# Prediction and location of malignant nodules and microcalcifications in mammography via Deep Learning

**DOI:** 10.1101/2022.10.11.22280939

**Authors:** David Coronado-Gutiérrez, Pablo Franco, Carlos López

## Abstract

**Objectives:** To propose a tool to detect and locate malignant nodules and microcalcifications in mammography and judge its potential as a screening tool.

**Methods:** In this institutional review board approved retrospective study we presented a new tool based on deep learning techniques to predict and locate lesions in mammograms, called quantusMM. 3,114 mammograms from 976 patients were collected from Onkologikoa (Instituto Oncológico de Kutxa) databases for this purpose: 1,248 images with malignant nodules, 736 with malignant microcalcifications and 1,131 without any suspicious findings. The proposed methods split the images in patches to be able to locate the lesions in the image. Then, these methods select the patches most likely to have a lesion based on the brightness values of the pixels. 80% of the selected patches (with the corresponding outcome) were used to train deep learning algorithms and the remaining 20% were used to test the performance to classify into malignant parts or control parts.

**Results:** The proposed methods obtain an area under the ROC curve (AUC) of 95.5% to predict malignant nodules using the patches, and 90.4% to predict malignant nodules into the whole images. To predict malignant microcalcifications the method obtains an AUC of 99.0% into patches and 90.0% into the whole images.

**Conclusions:** The proposed tool shows potential to predict and locate malignant nodules and microcalcification lesions in mammography. This new approach could help in the first screening of patients and also could greatly benefit radiologists to support decision making.

## Introduction

Breast cancer is the most common type of cancer among women and represents 15% of the total tumors worldwide [1]. Although research in this field has increased greatly in recent years, female breast cancer is still the fifth leading cause of death with more than 650,000 deaths per year [2]. To reduce the mortality rate, early diagnosis and proper assessment is crucial. Mammography is the most useful tool for general population screening but its performance to detect and diagnose breast cancer lesions highly depends on the radiologist expertise [3].

In recent years, the introduction of artificial intelligence methods in medical analysis has brought forth a potential revolution in computer-based interpretation of digital mammography [4]. Since the emergence of deep learning techniques and especially convolutional neural networks (CNN), many works have been published exploiting deep architectures in the field of breast cancer imaging [5]. In this way, CNN methods were extensively used in several studies for breast cancer classification [6], [7], nodules diagnosis [8], [9], microcalcification diagnosis [10], [11], tumor, nodules or microcalcifications segmentation [12], [13] and many others tasks.

In this study we proposed a new tool based on CNN methods to predict and locate malignant nodules and microcalcifications with a different approach. Firstly, we proposed to divide the entire images into patches as is done in other imaging modalities. This will allow us to analyze large resolution images with more accuracy and to more easily detect the location of the lesion in the breast. Secondly, we proposed to display the results in a similar way to the BIRADS system [14], a method used by radiologists to classify mammographic findings. Displaying the results based on a grade system as BIRADS will allow it to be more intuitive to clinicians and they will have more information of the possible severity of the lesion.

Therefore, the main objective of this study is to propose an innovative tool to detect and locate malignant nodules and microcalcifications in mammography and judge its potential as a screening tool.

## Materials and Methods

### Study design

This was an institutional review board approved retrospective study. The data was recoded in Onkologikoa, Instituto Oncológico de Kutxa (San Sebastain, Spain) between May 2008 and January 2021. All the acquired images were mammograms of craniocaudal (CC) or mediolateral oblique (MLO) views stored in DICOM (Digital Imaging and Communication in Medicine) format. The images were acquired by specialized breast-imaging radiologists during each patient examination with no prior guidelines. The protocol for collect the images was approved by the ethics committee of Osakidetza (Área Sanitaria de Gipuzkoa) under protocol identifier CUR-DIG-2020-01, with a waiver of the need to obtain informed consent.

A total of 3,114 mammograms from 976 patients were collected for this study. Three different breast cancer specialists manually diagnosed and delineated nodules and microcalcifications in all breasts images. To do this task, the clinicians used an online graphical user interface created by Transmural Biotech (Barcelona, Spain) [15]. Finally, from these 3,114 images, 1,248 images (666 patients) were breasts with diagnosed malignant nodules, 736 (378 patients) were breasts with diagnosed malignant microcalcifications and 1,131 (310 patients) were breasts without any suspicious findings.

### Methods

The goal of this study was to create a new tool based on deep learning techniques to predict and locate lesions in mammograms. This new approach was called quantusMM and it is formed by two independent modules: a detector of malignant nodules and a detector of malignant microcalcifications. Both modules first split the image in patches to be able to easily work with high resolution images and also to be able to give a prediction of the location of the lesion. Before that, the system homogenizes all the images by applying a normalization algorithm.

All the methods described in this section were developed using Matlab R2018a (MathWorks, Inc., Natick, MA, USA) programming language, except the “image normalization” subsection which was developed using Python 3.9 (Python Software Foundation, Wilmington, DE, USA).

#### Image normalization

To normalize the images that will be used in the prediction modules, the system first transforms all the DICOM images to “MONOCHROME2” format (Photometric Interpretation Attribute). Subsequently, the system applies a contrast enhancement with CLAHE algorithm [16] using OpenCV library [17]. The last step is to resize the images to 4096 pixels height, keeping the scaling for the width pixels.

#### Malignant nodules prediction

The nodules prediction module first splits all the processed images (breasts both with and without lesions) in patches and applies a candidate detection algorithm to discard the parts with less information. The detection algorithm is based on the intensity of the pixel values as we know that in mammograms all the nodules have high brightness values. Then, all the selected patches (with the corresponding outcome) are used to train and test a deep learning algorithm to try to predict malignant nodule lesions (Figure 1).

**Figure 1.**
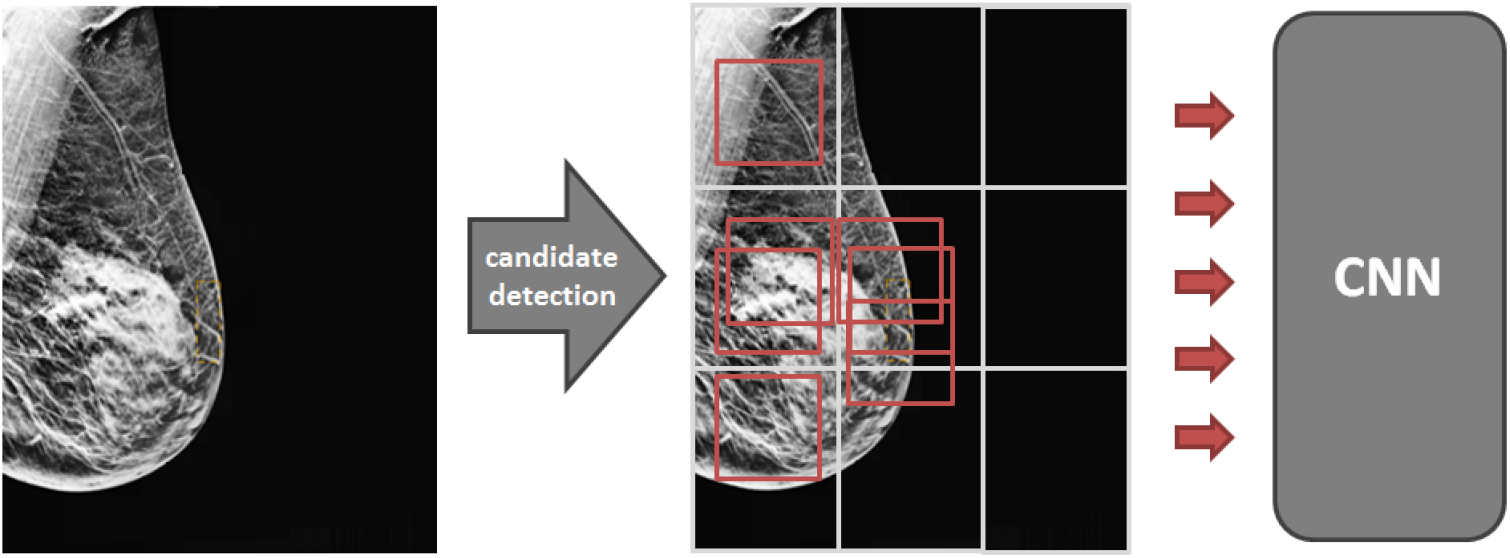
Outline of the steps that the initial mammography follows until the selected patches are used by the network to predict malignant nodules.

To obtain the patches, each image is swept with a window of 800 per 800 pixels in strides of 40 pixels (best values experienced). Then, each patch is assigned with an average brightness value considering the brightness values of all its pixels, giving more weight to the pixels in the center of the patch. Finally, each image is divided in 9 equal parts, and the 3 patches with higher brightness values of each part are selected, as long as they have a separation of 250 pixels (best value experienced) between them (Figure 1).

To obtain the outcome of all selected patches we assigned a patch as a “case” if the patch contained at least 80% of a malignant nodules lesion (previously assigned by the clinicians). On the other hand, we assigned a patch as a “control” if the patch contains 0% of a malignant nodule lesion. Both “controls” and “cases” patches can come from breasts without lesions as well as from breasts with diagnosed lesions.

Finally, the selected patches are divided into training and testing subset. The training patches trained the network of the CNN model, and the testing patches tested the generated model to observe the performance to classify into malignant parts or control parts. The CNN used was a ResNet50 [18], trained during 7 complete passes through the dataset (epochs) with batches of 32 samples (batch size). A Softmax loss function [19] and Adam optimization algorithm [20] was also applied. The method also used data augmentation algorithms to increase the amount of data to train the model, generating new patches by translating and rotating the original patches.

#### Malignant microcalcification prediction

In the microcalcifications module all the processed images are split in patches. Then, all the patches (with the corresponding outcome) with a minimum value of brightness are used to train and test a deep learning algorithm to try to predict malignant microcalcification lesions (Figure 2).

**Figure 2.**
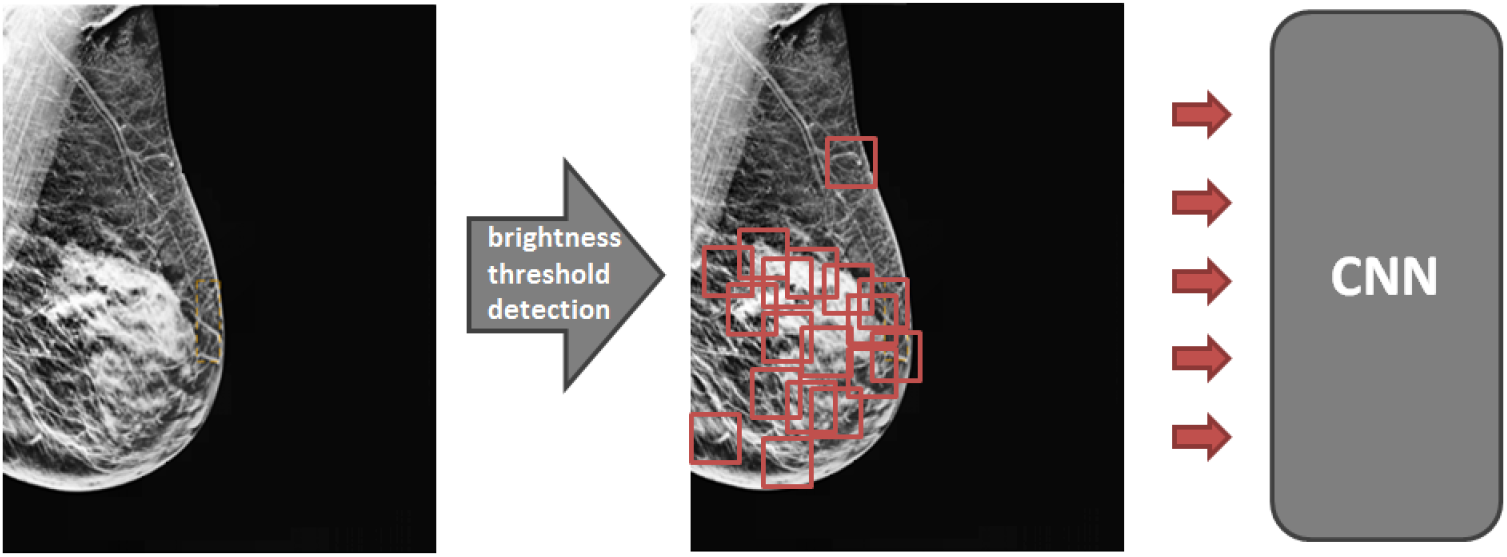
Outline of the steps that the initial mammography follows until the selected patches are used by the network to predict malignant microcalcifications.

To obtain the patches, each image is swept with a window of 400 per 400 pixels in strides of 200 pixels (best values experienced). Then, all the patches with at least one pixel with 90% of maximum brightness value (best values experienced) are selected.

To obtain the outcome of all selected patches we assigned a patch as a “case” if the patch contained the whole microcalcification lesion (previously assigned by the clinicians), otherwise the patch was considered a “control”. Both “controls” and “cases” patches can come from breasts without lesions as well as from breasts with diagnosed lesions.

Finally, the selected patches are divided into training and testing subset, same as in the case of nodules. The CNN used was also a ResNet50 [18], trained during 6 complete passes through the dataset (epochs) with batches of 32 samples (batch size). A Softmax loss function [19] and Adam optimization algorithm [20] was also applied. The method also used data augmentation algorithms to increase the amount of data to train the model, generating new patches by translating and rotating the original patches.

### Statistical analysis

We evaluated the performance of the two proposed modules with the same methodology. The selected patches of each module were randomly split 80% for the training subset and 20% for testing subset, preserving the prevalence of each class type and putting all the patches of the same image and patient in the same set.

After training the models, each testing subset was used to compute output scores for each patch and compared with the clinical outcome to obtain the receiver operator characteristic (ROC) curve and compute the area under the curve (AUC) for each module. Then, typical statistical metrics (sensitivity, specificity, etc.) were computed in different cut-off points, based on BIRADS classification which we will explain in the next section.

We also computed the ROC curve and statistical metrics by image, to observe the performance to predict lesions in whole images. To perform this, we assigned a “case” image if the image had one malignant lesion and a “control” image if the image did not have any lesions. We will do this both for the real outcome and for the patches predicted by the models.

### Online tool

The proposed tool quantusMM will be integrated into an online platform (www.quantusmm.org), making it available to any professional. The clinician could upload a mammogram and the platform will return a detailed report with the results obtained by the generated models in a few minutes. This report will show the probability of having a malignant lesion (nodule and microcalcification) based on BIRADS system [14] and also the location of the possible lesion in the breast (Figure 3).

**Figure 3.**
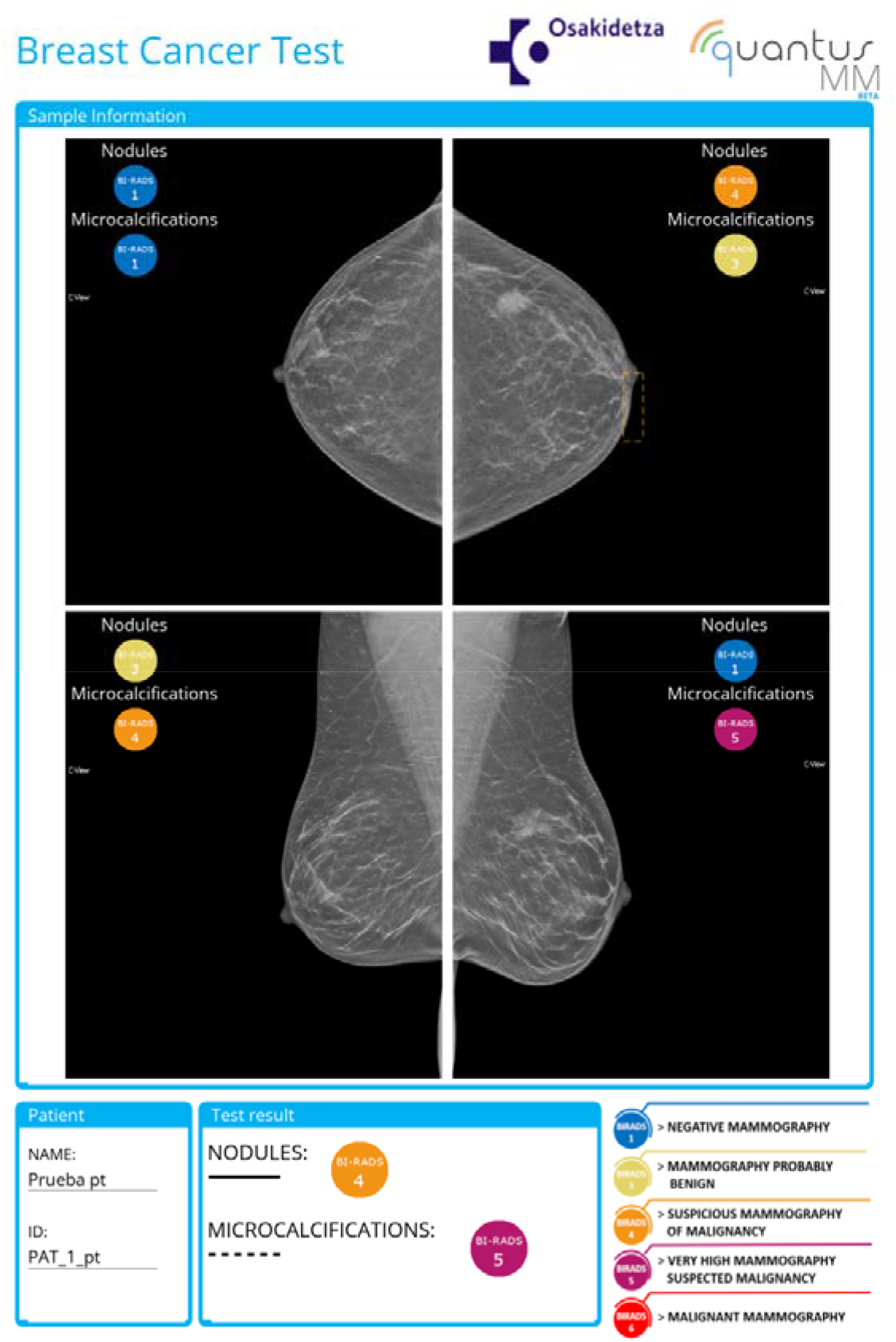
Example of a report obtained by the platform quantusMM.

To obtain the probability of lesion based on the BIRADS classification system, we set 4 different thresholds for the prediction values obtained with the CNN. BIRADS-6 means “malignant” and it is obtained for a specificity higher than 99%; BIRADS-5 means “very high suspicion of malignancy” and it is obtained for specificity between 95% to 99%; BIRADS-4 means “suspicion of malignancy” and it is obtained for a specificity lower than 95% and a sensitivity lower than 95%; BIRADS-3 means “probably benign” and it is obtained for a sensitivity between 95% and 99%; BIRADS-1 means “benign” and it is obtained for a sensitivity higher than 99%.

## Results

### Malignant nodules prediction

After the detection algorithm, 90,034 patches (85,690 controls and 4,344 cases) were selected to use in the nodules prediction module. Of these, 69,279 were used to train the network and the remaining 20,755 to observe the performance to classify into malignant or benignant. Figure 4 shows the ROC curve obtained by the proposed deep learning algorithm on the testing images, which obtained an AUC of 95.5%.

**Figure 4.**
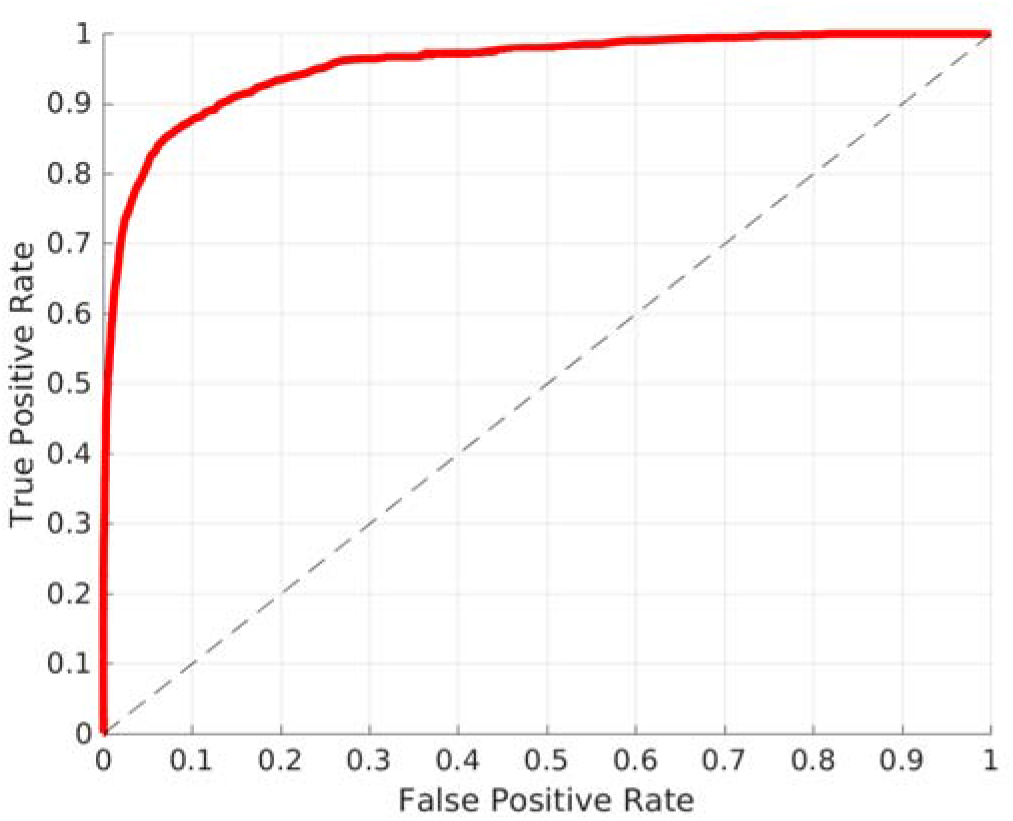
ROC curve of the nodules prediction module obtained for the patches test set.

**Figure 5.**
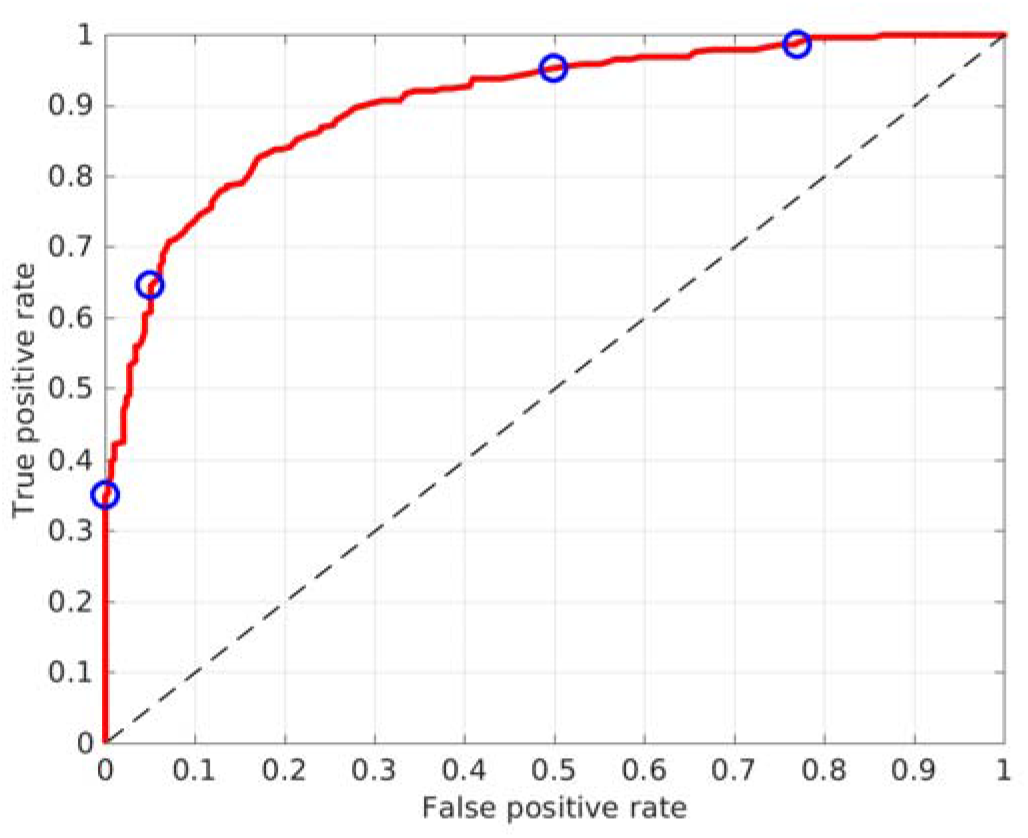
ROC curve of the nodules prediction module computed for whole images only. The blue circles point the different cut-offs depending on the BIRADS classification assigned (Table1).

On the other hand, if we compute the results only for whole images and not for patches, we would obtain the ROC curve of Figure 4, which obtained an AUC of 90.4%. Table 1 shows the detailed metric scores at several cut-off points of this ROC curve, chosen to later show the correct BIRADS classification.

**Table 1.** Statistical metrics obtained in the previous ROC curve (Figure 5) at 4 different cut-off points based on BIRADS classification [14]. (PPV = Positive Predictive Ratio; NPV = Negative Predictive Ratio).

|  | Accuracy | Sensitivity | Specificity | PPV | NPV |
| --- | --- | --- | --- | --- | --- |
| **BIRADS-6** | 68% | 37% | **99%** | 99% | 62% |
| **BIRADS-5** | 80% | 65% | **95%** | 92% | 73% |
| **BIRADS-4** | 73% | **95%** | 51% | 65% | 91% |
| **BIRADS-3** | 61% | **99%** | 23% | 56% | 95% |

### Malignant microcalcification prediction

After the detection algorithm, 67,846 patches (59,781 controls and 8,065 cases) were selected to use in the microcalcifications prediction module. Of these, 49,205 were used to train the network and the remaining 18,641 to observe the performance to classify into malignant or benignant. Figure 6 shows the ROC curve obtained by the proposed deep learning algorithm on the testing images, which obtained an AUC of 99.0%.

**Figure 6.**
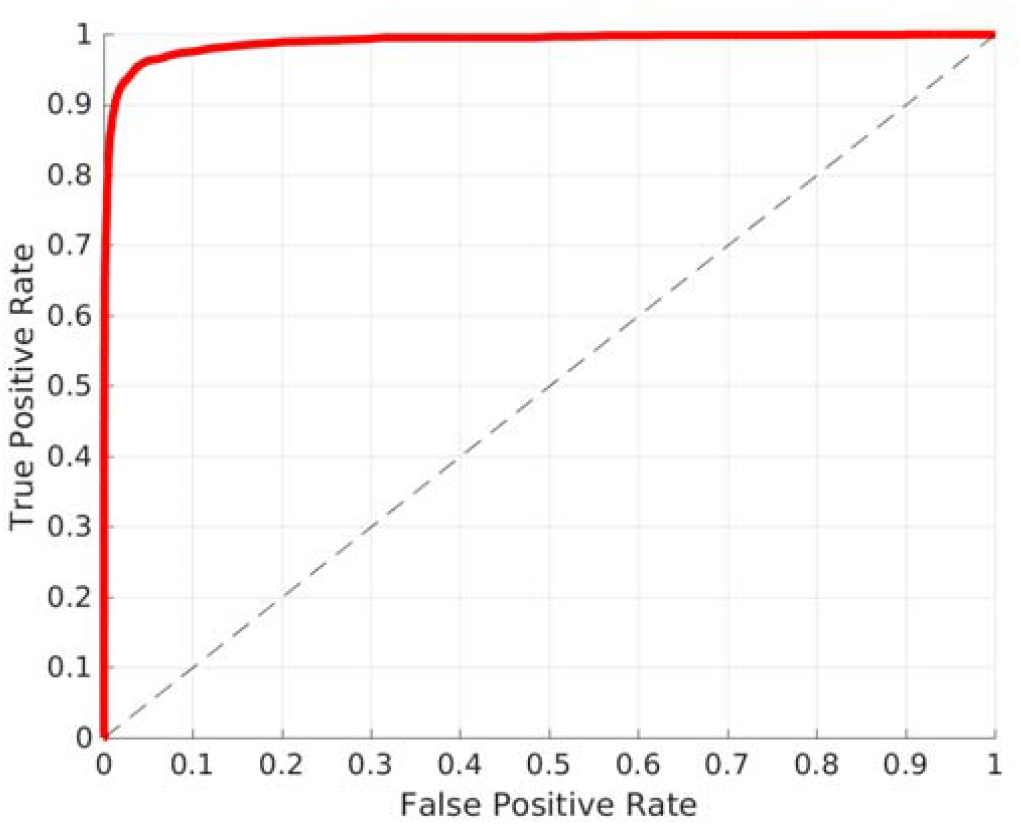
ROC curve of the microcalcifications prediction module obtained for the patches test set.

On the other hand, if we compute the results only for whole images and not for patches, we would obtain the ROC curve of Figure 7, which obtained an AUC of 90.0%. Table 2 shows the detailed metric scores at several cut-off points of this ROC curve, chosen to later show the correct BIRADS classification.

**Table 2.** Statistical metrics obtained in the previous ROC curve (Figure 7) at 4 different cut-off points based on BIRADS classification [14]. (PPV = Positive Predictive Ratio; NPV = Negative Predictive Ratio).

|  | Accuracy | Sensitivity | Specificity | PPV | NPV |
| --- | --- | --- | --- | --- | --- |
| **BIRADS-6** | 77% | 42% | **99%** | 99% | 73% |
| **BIRADS-5** | 84% | 66% | **95%** | 89% | 81% |
| **BIRADS-4** | 68% | **95%** | 52% | 56% | 94% |
| **BIRADS-3** | 40% | **99%** | 3% | 39% | 89% |

**Figure 7.**
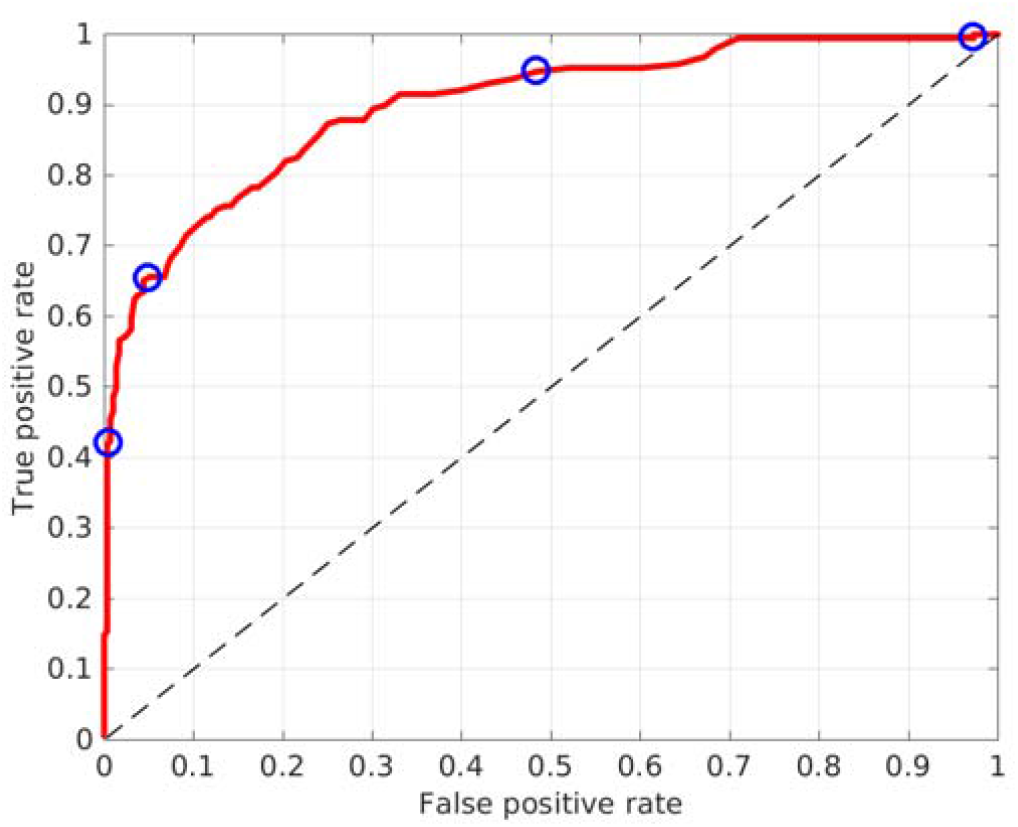
ROC curve of the microcalcifications prediction module computed for whole images only. The blue circles point the different cut-offs depending on the BIRADS classification assigned (Table2).

## Discussion

We built a new tool called quantusMM based on state-of-the-art deep learning techniques to automatically predict malignant nodules and microcalcifications in mammography and place them in the image. The proposed techniques were evaluated on a database of 3,114 images selected for the purpose of this study.

Results showed that the proposed tool can effectively predict malignant nodules and microcalcifications on mammograms. The obtained ROC curves reached an AUC of 90.4% and 90.0% to predict nodules and microcalcifications in whole images, respectively. These results open the door to its potential use as a screening tool on patients with breast cancer risk factors.

The statistical metrics also showed a good performance with the different cut-offs points selected on the previous ROC curves. These cut-off points represent the different BIRADS classifications which give the platform tool, depending on the prediction result of each image. Giving the results not as a binary system (malignant / non-malignant) but as a degree system similar to BIRADS, allows to give more information to radiologists and could greatly benefit them to support decision making.

On the other hand, the division of the images in patches allows us to easily work with mammograms which are high resolution images. The patches division also allows us to predict the lesion location in the breast, even if the lesions are small like the microcalcifications. The results obtained in lesion prediction in patches are better than those obtained with the whole images. The AUC obtained was 95.5% and 99.0% for nodules and microcalcifications, respectively. This would indicate that the precision to detect malignant patches is high, and therefore, the subsequent precision to lesion location in the whole image will be good.

This study has strengths and limitations. The main strength is that the collected database is a real database acquired from clinical practice, without any specific acquisition protocol for this study. This gives robustness to the proposed tool and indicates that it could work well with new images from clinical practice. One limitation of the study is that the small size of the data test set. It is recommended to confirm these results in larger multicenter studies to corroborate the trend of the results. Another limitation is that we only used images of healthy breasts or images with nodules or microcalcification lesions. Any other lesion has been introduced in the training method, which could worsen the results if an image with a different lesion was tested.

To conclude, the proposed tool based on Deep Learning algorithms shows potential to predict and locate malignant nodules and microcalcification lesions in mammography. This new approach could help in the first screening of patients and also could greatly benefit radiologists to support decision making. A large clinical study is recommended to confirm these results.

## Data Availability

All data associated with this study is property of Osakidetza (Servicio Vasco de Salud).

## Data and materials availability

All data associated with this study is property of Osakidetza (Servicio Vasco de Salud). The developed AI method is available at www.quantusmm.org.

